# Prevalence of childhood cough in epidemiological studies: does it matter which question we use?

**DOI:** 10.1101/2021.03.29.21254350

**Authors:** Maria Christina Mallet, Rebeca Mozun, Cristina Ardura-Garcia, Philipp Latzin, Alexander Moeller, Claudia E. Kuehni

**Author notes:** Corresponding author: Prof. Dr. med Claudia E. Kuehni, Institute of Social and Preventive Medicine, University of Bern, Mittelstrasse 43, 3012, Bern, Switzerland.

## Abstract

We assessed how prevalence estimates of cough in 6-17-year-olds vary depending on the question asked in the population-based *Luftibus in the school* (LUIS) study. 3427 parents answered three different questions on cough. The prevalence of parent-reported cough varied substantially depending on the question: 25% of parents reported cough without a cold, 11% dry night cough and 5% that their child coughs more than other children. There was only partial overlap with 3% answering yes to all questions. This suggests that the exact question used to assess cough strongly affects prevalence estimates and must be taken into account when comparing studies.

## INTRODUCTION

Epidemiological studies often estimate prevalence of cough through parental questionnaires. Cough is common in children and occurs as a physiological response during upper respiratory tract infections, but frequent cough in the absence of an airway tract infection is typical for chronic respiratory diseases such as asthma.[1] Different research networks have developed specific questions to identify children with cough that exceeds the normal expected physiological occurrence. The American Thoracic Society (ATS) questionnaire asks if the child has “a cough even without having a cold”.[2] The International Study of Asthma and Allergies in Childhood (ISAAC) asks for “a dry cough at night, apart from a cough associated with a cold or a chest infection”[3] and a questionnaire developed in Southampton and used in several studies inquires if the child “coughs more than other children”.[4]

Most epidemiological studies conducted in the field of child respiratory health have only included one of these cough questions basing the prevalence of cough on this single question.[5-7] This has made it impossible to distinguish whether differences in prevalence of cough between studies reflect regional variations, differences in study populations, varying exposure to environmental risk factors, or if they result only from differences in the wording of the question.

We used data from a large population-based study that included all three questions on cough in the same questionnaire, to assess how prevalence of parent-reported cough in children varies depending on the question that is used.

## METHODS

We analysed data from *LuftiBus in the School* (LUIS), a cross-sectional population-based study on respiratory health in schoolchildren, conducted in the canton of Zurich, Switzerland, in 2013-2016.[8] Schools were dispersed across the entire canton and the proportion of rural and urban schools and the area-based socioeconomic index of participants was typical for the region.[8] Parents provided informed consent and completed a questionnaire on respiratory symptoms of their child, including three different questions about cough: *“Does your child have a cough even without having a cold?”* (cough without a cold)[2], *“In the last 12 months has your child had a dry cough at night, apart from a cough associated with a cold or a chest infection?”* (dry night cough)[3] and *“Do you think your child coughs more than other children?”* (cough more than others).[4] We compared the prevalence of cough as assessed using the three questions and constructed a Venn diagram to describe the degree of overlap between parents’ answers. We also stratified the analyses for factors that could affect the prevalence of cough that are: current wheeze (as a proxy for asthma), sex, and age group. We also assessed the duration of cough continuing for more than three weeks and more than two months.

## RESULTS

Among 3870 participants from 37 schools, 3427 had completed parental questionnaires and informed consent, and were included in the analysis. Median age was 13 years (range 6-17), 50% (1723) were girls and 8% (280) reported current wheeze (during the past 12 months).

The prevalence of cough differed substantially depending on the question asked. 872 (25%, 95% CI 24 – 27%) parents reported cough without a cold, 390 (11%, 95% CI 10 – 12%) dry night cough and 158 (5%, 95% CI 4 – 5%) answered that their child coughs more than other children.

There was only partial overlap between the responses to the three cough questions (Figure 1A): 183 (5%) reported cough without a cold and dry night cough, but not coughing more than others; 88 (3%) answered yes to all three questions and only 13 (<1%) reported that their child coughs more than other children without reporting also cough without a cold or dry night cough.

**Figure 1:**
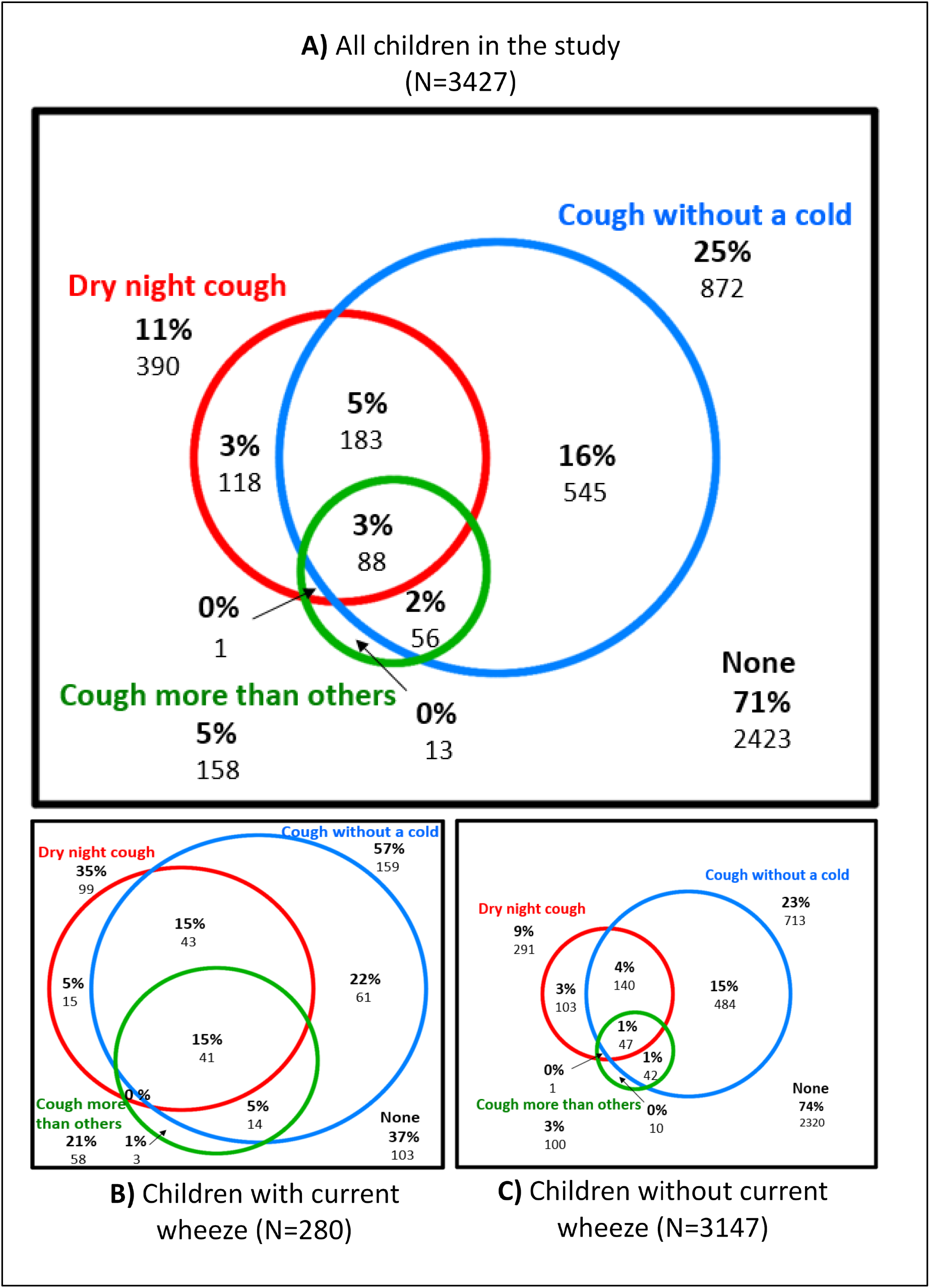
Venn diagram showing the overlap between parents’ answers to three different questions on cough in (A) the whole study population (N=3427), (B) children with current wheeze (N=280) and (C) children without current wheeze (N=3147)

For children with current wheeze, parents reported a higher prevalence of cough, for all questions. Only 37% of the 280 children with current wheeze had no parental report of cough, while this proportion was 74% for the 3147 children without current wheeze. Also the overlap across positive answers to the three cough questions was greater in children with current wheeze (Figure 1B) than in children without wheeze (Figure 1C). In children with wheeze, 15% of parents affirmed all three questions on cough; this proportion was only 1% in children without wheeze. The overlap did not vary substantially when we stratified the population by age or sex.

A high proportion of parents who reported that their child coughed more than other children also reported episodes of cough lasting more than three weeks (51%) or more than two months (11%). These proportions were much lower among parents who reported cough without a cold (22% and 3%) or dry night cough (29% and 5% respectively).

## DISCUSSION

This cross-sectional study suggests that parents’ answers to different questions on cough vary substantially, with prevalence of cough ranging between 5% and 25% depending on the question used. There was only partial overlap between the answers.

Few studies reporting on cough in population-based studies used more than one question.[9, 10] The Leicester cohort study (LRC) is one study that included all three. Compared to LUIS, prevalence of cough was twice as high in the LRC, with 47% of 10-13-year olds reporting cough without a cold, 22% dry night cough and 10% cough more than others perhaps due to the higher prevalence of asthma (21% versus 9%). However, differences between the cough questions were comparable.[10]

The question whether the child coughs more than other children was least often answered in the affirmative. For these children, parents usually reported also cough without a cold and dry night cough and longer cough episodes. Therefore, this question might be most helpful to identify children with more severe cough caused by airway diseases such as asthma.

Our findings have implications for epidemiological studies in children. The wording of the question used to assess cough strongly affects prevalence estimates, resulting in five-fold differences (5% to 25%). This is much more than the expected effects of environmental exposures such as air pollution or tobacco smoke. Only studies using exactly the same questions should be compared.

## Data Availability

Study collaborators and other researchers can obtain datasets for analysis if a detailed concept sheet is presented for the planned analyses and approved by the principal investigators (AM, PL and CK).

## Acknowledgements

We would like to thank the LUIS study group and all the parents and children who participated in the study. We also thank Dr Eva Pedersen for her contribution in the discussions and Daria Berger for her contribution in editing.

## The Luftibus in the school study group

Alexander Moeller, Jakob Usemann (Division of Respiratory Medicine, University Children’s Hospital Zurich and Childhood Research Center, University of Zurich, Switzerland); Philipp Latzin, Florian Singer and Johanna Kurz (Paediatric Respiratory Medicine, Children’s University Hospital of Bern, University of Bern, Switzerland); Claudia E. Kuehni, Rebeca Mozun, Cristina Ardura-Garcia, Myrofora Goutaki, Eva S.L. Pedersen and Maria Christina Mallet (Institute of Social and Preventive Medicine, University of Bern, Switzerland); Kees de Hoogh (Swiss Tropical and Public Health Institute, Basel, Switzerland).

## Contributors

AM, CK and PL conceptualised and designed the LUIS study. AM supervised data collection. MCM, CK, RM and CAG conceptualised this analysis. MCM analysed the data and drafted the manuscript. CK, RM, CAG, AM and PL provided input for interpretation of findings. All authors critically reviewed and approved the final manuscript.

## Funding

Lunge Zürich, Switzerland, funded the study set-up, development, and data collection with a grant to Alexander Moeller. Analysis has been supported by a grant of the Swiss National Science Foundation (320030_182628) to Claudia Kuehni.

## Competing Interests

Dr. Latzin reports personal fees from Gilead, Novartis, OM pharma, Polyphor, Roche, Santhera, Schwabe, Vertex, Vifor, Zambon and grants from Vertex, all outside the submitted work. Dr. Moeller reports personal fees from Vertex and OM Pharma, outside the submitted work.

## Patient consent for publication

Not required

## Ethics approval

Approved by the ethics committee of the canton of Zurich (KEK-ZH-Nr: 2014-0491).

## Notes

### Funding Statement

Lunge Zurich, Switzerland, funded the study set-up, development, and data collection with a grant to Alexander Moeller. Analysis has been supported by a grant of the Swiss National Science Foundation (320030_182628) to Claudia Kuehni.

### Author Declarations

Approved by the ethics committee of the canton of Zurich (KEK-ZH-Nr: 2014-0491).

